# Does the Health System Model Shape Prevention? Evidence from 22 OECD Countries (2004-2023)

**DOI:** 10.64898/2026.03.17.26348034

**Authors:** Pietro Marraffa, Lorenzo Marega, Gianfranco Politano, Maria Michela Gianino

**Author notes:** Correspondence concerning this article should be addressed to Marega, Lorenzo.

## Abstract

In an era in which population ageing, rising healthcare costs and growing global health challenges are pressing global issues, the main aim of our article is to analyze trends in preventive care expenditures from 2004 to 2023 in 22 European countries, examining whether specific health systems are associated with different time trends in preventive care expenditures over the considered time. Although there are few studies investigating this issue adopting the standard tripartite classification, to our knowledge, this is the first study to explore the topic using the latest classification of healthcare systems proposed by Böhm.

We performed a time trend analysis using secondary data from 22 European OECD countries during a twenty-year period (2004-2023); in addition, a hierarchical semi-log polynomial mixed-effects regression analysis has been performed, including annual country-level % preventive expenditures in association with the three structural dimensions — regulation, financing and provision — according to Böhm’s classification as explanatory variables.

Our results indicate that, in terms of compound annual rate, most countries exhibited an increase in % of preventive expenditures (between 0.2% and 3.7%), while seven countries denounced a decrease (between −6.3% and −0.2%) during the considered period. The regression analysis shows that the trend of % preventive expenditures did not differ in two of the three dimensions under study: financing and provision. In contrast, in countries with statal regulation, the curvilinear trend was more pronounced than in countries with statal regulation (b=0.0035; 95% CI= 0.0013, 0.0057).

In conclusion, there is no correlation between the type of healthcare system and the share of expenditure allocated to prevention activities in the countries analysed; a resulting implication is that investment in prevention is not intrinsically determined by the organisational structure of the healthcare system, but responds to external factors.

**Key questions:** *What is already known on this topic?:* Preventive care represents a relatively small share of total health expenditure in most OECD countries, despite its recognized importance in addressing public health issues. Previous studies attempted to explore cross-country differences in preventive spending and the potential role of healthcare system organization, often using traditional classifications (e.g., Beveridge or Bismarck). However, evidence remains limited and no studies have examined long-term trends using current multidimensional classifications of healthcare systems.

*What does this study add?:* By analyzing trends in preventive care expenditures over a twenty-year period across 22 European OECD, our study showed trends in the share of spending on prevention were largely independent of the structural characteristics of healthcare systems. Among the analyzed dimensions, only the regulation showed a more pronounced curvilinear trend in countries with societal regulation.

*How this study might affect research, practice or policy?:* Since the findings suggest that investment in prevention may depend more on contextual factors such as political priorities and public health strategies rather than structural characteristics of healthcare systems, policymakers should therefore promote prevention through targeted policy commitment instead of relying on health system design alone.

## INTRODUCTION

In an era of aging populations, rising healthcare costs, and growing global health challenges, the need for health promotion and disease prevention has never been more critical: for this very reason, stepping up work on preventive healthcare is one of the key European Commission’s health Political priorities for the period 2024-2029. (1).

While there is a consensus on the prevention of the diseases as the most valuable tool in order to reduce the burden of disease and associated risk factors (2), there is no homogeneity in the behavior regarding the expenditures on prevention by different countries, besides some governments invest more than others in public health services, as is suggested by the literature on health care systems (3–6). However, the few studies that investigated the relationship between preventive care expenditures and the type of healthcare system found that the latter is a determinant of preventive care investments, adopting the classic ideal-typical distinction between national health insurance, societal health insurance and the private healthcare system (3,4).

The breakdown of healthcare systems based on these three ideal types can be considered the standard tripartite classification (7) and, to our knowledge, the only exception consisted in a study that has adopted a different classification of healthcare systems conducted by Trein (4). The study hypothesized the relationship between preventive and curative fields depends on the healthcare system in a country, that is, the distinct forms of governance in three dimensions: regulation, financing, and provision of healthcare (8).

Over the past few decades, healthcare expenditures have become a key indicator of governments’ commitment to the well-being of their populations. Yet, performances on preventive care expenditures is a field that needs to be studied in greater detail. The literature documented a lack of funding for preventive care (3) and cuts to public health budgets, especially, after the economic crises (9) but very little studies on the trend of preventive care spending over the time and few comparative research has been conducted across countries (3,9,10).

In order to contribute more broadly to the literature on preventive care expenditures and address some shortcomings, this article aims to analyze trends in preventive care expenditures from 2004 to 2023 in 22 European countries with different healthcare systems, and examine whether specific health systems are associated with different time trends in preventive care expenditures.

## METHODS

A time trend analysis was performed using secondary data from 22 European OECD countries during a twenty-year period (2004-2023). The countries included in the study were: Austria, Belgium, Czech Republic, Denmark, Estonia, Finland, France, Germany, Hungary, Iceland, Ireland, Italy, Luxembourg, Netherlands, Norway, Poland, Portugal, Slovak Republic, Spain, Sweden, Switzerland and the United Kingdom. These countries and years were chosen based on the availability of data. We obtained official data from the Organization for Economic Co-operation and Development (OECD). Percentage preventive care expenditure to total healthcare expenditures was considered, calculated from values expressed in Euros, PPP converted, 2020.

Over the time, many proposals have been put forward to classify healthcare systems. We adopted the typology that was presented by Rothgang and Wendt (11) and modified by Böhm (8) because it attempts a deductive construction of healthcare system types and allows for a more precise classification of healthcare systems.

According to Böhm, each health care system is identified by three dimensions that are not entirely independent of each other but follow a clear order: the regulation dimension is first, followed by the financing dimension and finally service provision. In every dimension, three actors can play a role: state, societal or private actors (see Additional file 1 for a summary of Böhm’s classification) (8). Consequently, healthcare systems in OECD countries can be grouped under five main models. In addition to the time trend analysis, we performed a regression analysis including the three structural dimensions — regulation, financing and provision— according to Böhm’s classification as explanatory variables in order to explore their association with the share of preventive expenditure at the individual country level.

### Statistical analysis

We performed a hierarchical semi-log polynomial mixed-effects regression analysis of annual country-level % preventive expenditures, specifying random intercepts and random time slopes to account for unobserved heterogeneity across countries and the repeated-measures (country–year) structure of the panel (12). Observations were restricted to years ≥2014, and time was re-parameterized as a re-centered linear index (year_linear = year − (min(year) − 1)) with a quadratic term (year_linear^2) to capture curvature while improving numerical stability. The dependent variable was log-transformed (log(% preventive expenditures)) to improve model fit under convex/exponential time patterns and to support interpretation on a multiplicative scale. Fixed effects included year_linear, year_linear^2, and three categorical health system dimensions (regulation, financing, provision), each coded with “Statal” as the reference level, along with interactions between each dimension and both time terms to assess whether health system typologies were associated with different % preventive expenditures trajectories over time. The random-effects structure included a country-specific random intercept and country-specific random slopes for both year_linear and year_linear^2. Models were estimated by maximum likelihood using lmer from the lme4 package (12).

Because inference in mixed-effects models can be sensitive with a limited number of higher-level units, we quantified uncertainty for fixed-effect parameters using a model-based parametric bootstrap with 1000 replicates, implemented via bootMer, extracting the fixed effects at each replicate to obtain bootstrap standard errors and normal-approximation 95% confidence intervals and two-sided p-values (12,13).

Random-effects variability was summarized using the estimated standard deviations of the country-level random intercept and random slopes, alongside the residual (country–year) standard deviation from the fitted model. To support interpretation of between-group differences, we derived estimated marginal means (overall level differences) and estimated marginal trends (differences in slopes with respect to year_linear) for each health system dimension using the emmeans framework, including global joint tests and pairwise contrasts (14). For visualization and reporting, fitted values and confidence limits computed on the log scale were back-transformed to the original % preventive expenditures scale using the exponential function. All analyses were conducted in R version 4.3.1 (2023-06-16) using the major packages lme4 (model estimation and parametric bootstrap), emmeans (marginal means and trends), dplyr/tidyr/purrr (data management), and ggplot2 (visualization) (12,14–18).

## RESULTS

Figure 1 displays the % of preventive expenditure in 22 European countries examined. Most countries exhibited an increase in % of preventive expenditures during 2004-2023, and seven countries (Hungary, Ireland, Island, Poland, Portugal, Slovak Republic, Switzerland) denounced a decrease during the period studied. Figure 1 highlights that there is a wide range of initial and the final % preventive expenditures and highlights a disharmonious trend whereby countries did not maintain the same position in the final ranking compared to the initial one. All countries had a pick in 2021, after COVID.

**Figure 1.**
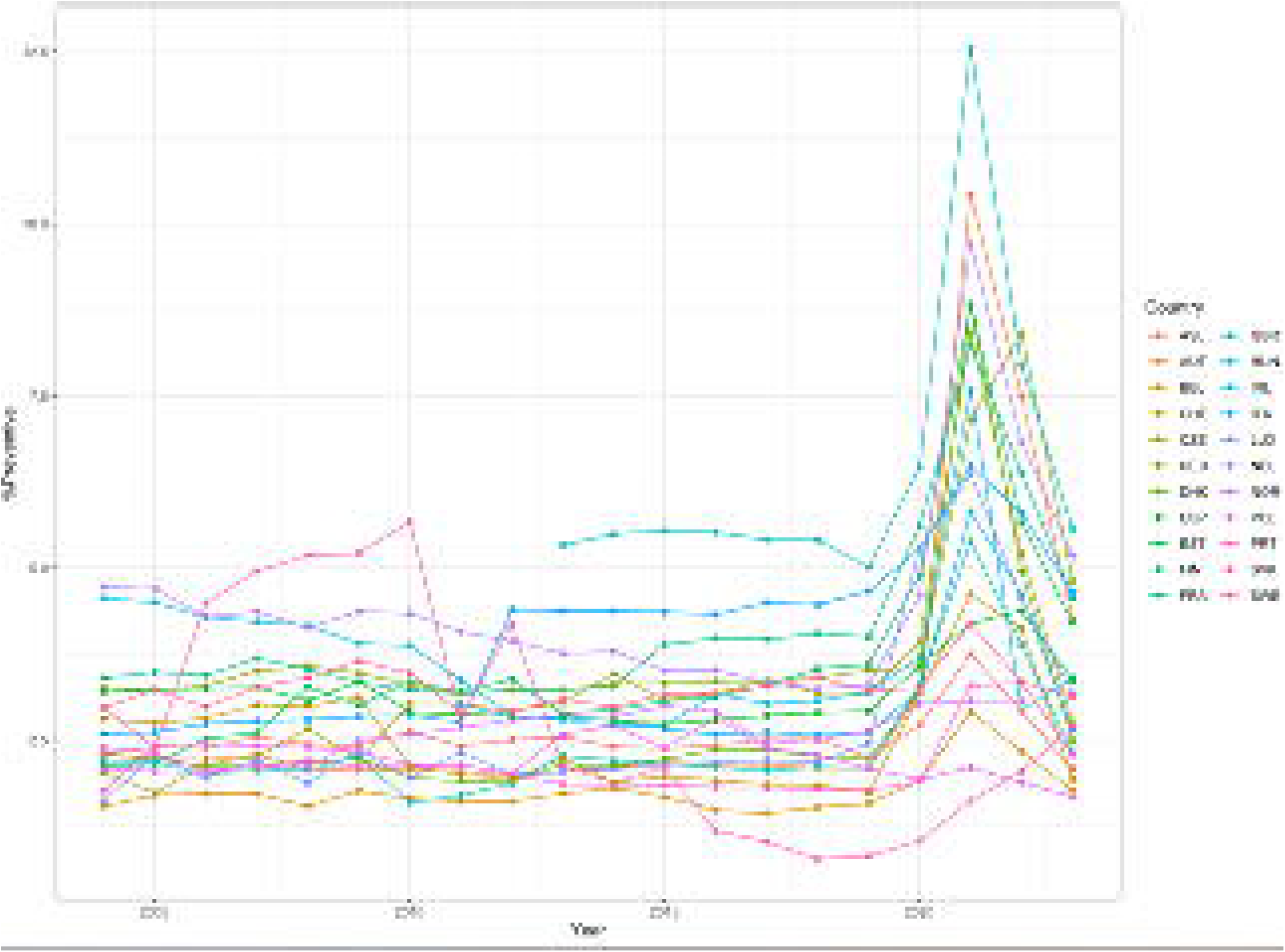
Percentage of preventive expenditure 2004-2013 in 22 European countries. Abbreviations: ASL Iceland; AUT, Austria; BEL, Belgium; CHE, Switzerland; CZE, Czech Republic; DEU Germany; DNK, Denmark; ESP, Spain; EST, Estonia; FIN, Finland; FRA, France; GBR, United Kingdom; HUN, Hungary; IRL, Ireland; ITA, Italy; LUX, Luxembourg; NDL, Netherlands; NOR, Norway; POL, Poland; PRT, Portugal; SVK, Slovak Republic; SWE, Sweden.

Table 1 presents the compound annual rate for each country. In ten-year period the % preventive expenditure grows at a compound annual rate between 0.2% and 3.7% in most countries. Conversely, in seven countries there is a decrease at a compound annual rate between −6.3% and −0.2% and, with the exception of Polonia, confirmed the decreasing trend already recorded in the period 2004-2019. All countries saw a sharp increase in the COVID period (2019-2021) to reduce the % of preventive expenditures in the following two years (2022-2023).

**Table 1.**
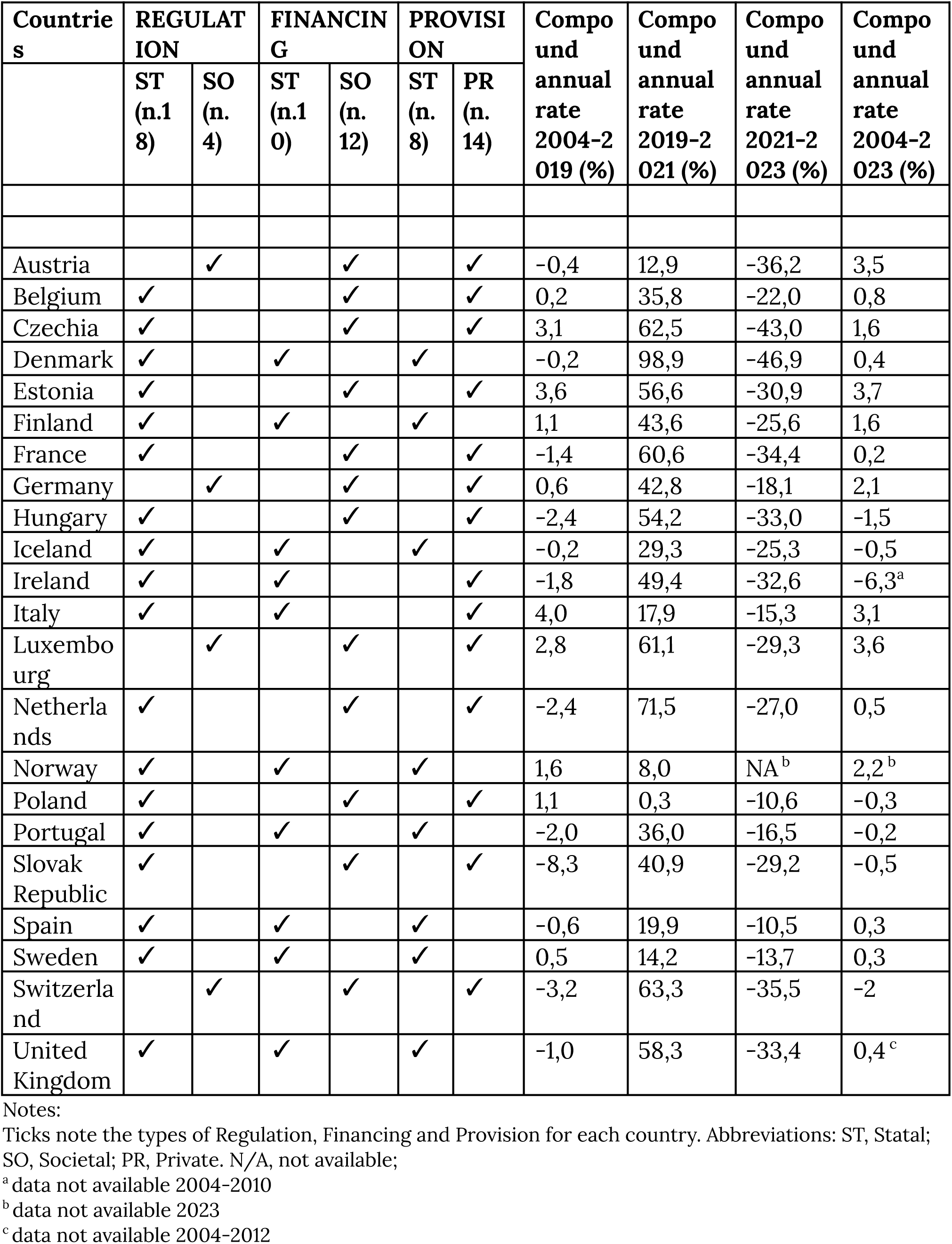
Compound annual rate of preventive expenditure (%) and types of Regulation, Financinfìg and Provision for each country.

| Countries | REGULATION |  | FINANCING |  | PROVISION |  | Compound annual rate 2004-2019 (%) | Compound annual rate 2019-2021 (%) | Compound annual rate 2021-2023 (%) | Compound annual rate 2004-2023 (%) |
| --- | --- | --- | --- | --- | --- | --- | --- | --- | --- | --- |
|  | ST (n.18) | SO (n.4) | ST (n.10) | SO (n.12) | ST (n.8) | PR (n.14) |  |  |  |  |
| Austria |  | ✓ |  | ✓ |  | ✓ | -0,4 | 12,9 | -36,2 | 3,5 |
| Belgium | ✓ |  |  | ✓ |  | ✓ | 0,2 | 35,8 | -22,0 | 0,8 |
| Czechia | ✓ |  |  | ✓ |  | ✓ | 3,1 | 62,5 | -43,0 | 1,6 |
| Denmark | ✓ |  | ✓ |  | ✓ |  | -0,2 | 98,9 | -46,9 | 0,4 |
| Estonia | ✓ |  |  | ✓ |  | ✓ | 3,6 | 56,6 | -30,9 | 3,7 |
| Finland | ✓ |  | ✓ |  | ✓ |  | 1,1 | 43,6 | -25,6 | 1,6 |
| France | ✓ |  |  | ✓ |  | ✓ | -1,4 | 60,6 | -34,4 | 0,2 |
| Germany |  | ✓ |  | ✓ |  | ✓ | 0,6 | 42,8 | -18,1 | 2,1 |
| Hungary | ✓ |  |  | ✓ |  | ✓ | -2,4 | 54,2 | -33,0 | -1,5 |
| Iceland | ✓ |  | ✓ |  | ✓ |  | -0,2 | 29,3 | -25,3 | -0,5 |
| Ireland | ✓ |  | ✓ |  |  | ✓ | -1,8 | 49,4 | -32,6 | -6,3<sup>a</sup> |
| Italy | ✓ |  | ✓ |  |  | ✓ | 4,0 | 17,9 | -15,3 | 3,1 |
| Luxembourg |  | ✓ |  | ✓ |  | ✓ | 2,8 | 61,1 | -29,3 | 3,6 |
| Netherlands | ✓ |  |  | ✓ |  | ✓ | -2,4 | 71,5 | -27,0 | 0,5 |
| Norway | ✓ |  | ✓ |  | ✓ |  | 1,6 | 8,0 | NA<sup>b</sup> | 2,2<sup>b</sup> |
| Poland | ✓ |  |  | ✓ |  | ✓ | 1,1 | 0,3 | -10,6 | -0,3 |
| Portugal | ✓ |  | ✓ |  | ✓ |  | -2,0 | 36,0 | -16,5 | -0,2 |
| Slovak Republic | ✓ |  |  | ✓ |  | ✓ | -8,3 | 40,9 | -29,2 | -0,5 |
| Spain | ✓ |  | ✓ |  | ✓ |  | -0,6 | 19,9 | -10,5 | 0,3 |
| Sweden | ✓ |  | ✓ |  | ✓ |  | 0,5 | 14,2 | -13,7 | 0,3 |
| Switzerland |  | ✓ |  | ✓ |  | ✓ | -3,2 | 63,3 | -35,5 | -2 |
| United Kingdom | ✓ |  | ✓ |  | ✓ |  | -1,0 | 58,3 | -33,4 | 0,4<sup>c</sup> |

Notes:
Ticks note the types of Regulation, Financing and Provision for each country. Abbreviations: ST, Statal; SO, Societal; PR, Private. N/A, not available;
<sup>a</sup> data not available 2004-2010
<sup>b</sup> data not available 2023
<sup>c</sup> data not available 2004-2012

Table 1 displays the classification of each healthcare system according to the three dimensions too.

Eight countries had statal regulation, financing and provision, 8 had statal regulation, societal financing, private provision; four countries had societal regulation and financing with private provision; two had statal regulation and financing with private provision.

Table 2 displays the results of regression analysis. The trend of % preventive expenditures did not differ in two of the three dimensions under study: financing and provision. In contrast, in countries with statal regulation, the curvilinear trend was more pronounced than in countries with statal regulation (b=0.0035; 95% CI= 0.0013, 0.0057).

**Table 2.** Results of hierarchical semi-log polynomial mixed-effects regression models on % preventive expenditure in 22 European countries 2004-2023.

| Variable | Regression coefficient | bootstrap standard error | p | Normal based 95% CI |  |
| --- | --- | --- | --- | --- | --- |
| (Intercept) | 1.055 | 0.0989 | 0 | 0.8612 | 1.249 |
| Year | -0.0216 | 0.0154 | 0.1596 | -0.0517 | 0.0085 |
| Year<sup>2</sup> | 0.0019 | <0,001 | 0.0044 | <0,001 | 0.0032 |
| Societal Regulation (ref. Statal) | 0.0383 | 0.1596 | 0.8103 | -0.2745 | 0.3512 |
| Societal Financing (ref. Statal) | 0.2864 | 0.2765 | 0.3002 | -0.2555 | 0.8283 |
| Private Provision (ref. Statal) | -0.2501 | 0.2699 | 0.3542 | -0.7791 | 0.2789 |
| Societal Regulation xYear | -0.0509 | 0.0261 | 0.051 | -0.102 | <0,001 |
| Societal Regulation xYear<sup>2</sup> | 0.0035 | 0.0011 | 0.0021 | 0.0013 | 0.0057 |
| Societal Financing x Year | -0.0688 | 0.042 | 0.1014 | -0.151 | 0.0135 |
| Societal Financing x Year<sup>2</sup> | 0.0018 | 0.0017 | 0.2853 | -0.0015 | 0.0052 |
| Private Provision x Year | 0.0571 | 0.0409 | 0.1626 | -0.0231 | 0.1373 |
| Private Provision x Year<sup>2</sup> | -0.0019 | 0.0017 | 0.2572 | -0.0052 | 0.0014 |

Abbreviations: 95% CI, 95% confidence interval; SD, standard deviation;
Notes: The quadratic term, year<sup>2</sup>, indicates the presence of a curvilinear (or nonlinear, U-shaped) trend over time.
The interaction terms (marked with the sign “x”) indicate how much year and year<sup>2</sup> are different for different health care system types. So, year and year<sup>2</sup> represent the linear and quadratic slope when regulation, financing and provision are issued by public actors, i.e., the reference category for each dimension.

Results of regression analysis are illustrated in Figure 2 where % preventive expenditures are stratified by dimension and plotted for each year. Only countries with societal regulation had a pronounced speed up in the increase of % preventive expenditure, and starting from the same level as countries with statal regulation, they likely best used the room to increase the % of spending during the study period.

**Figure 2.**
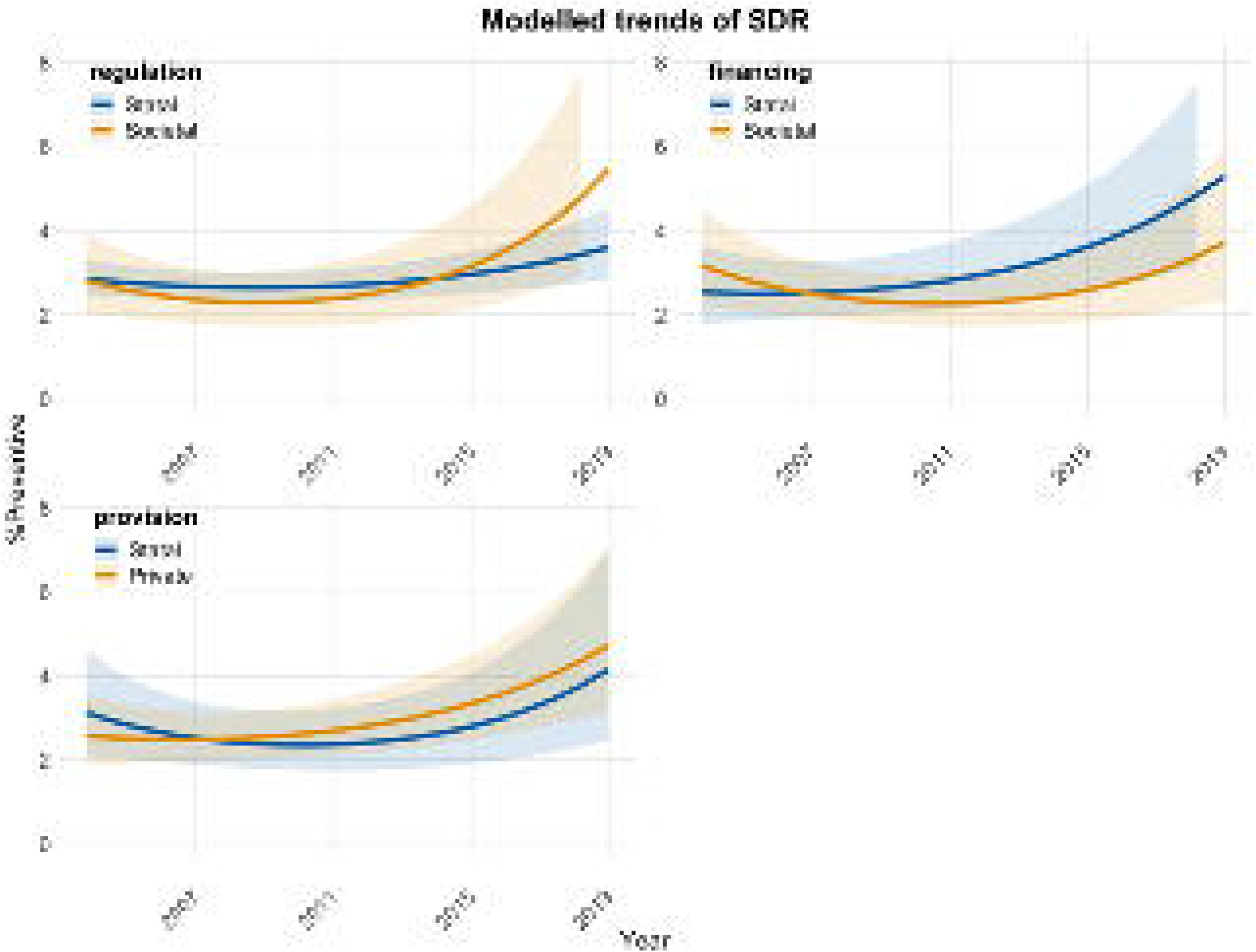
Modelled Trends Plot of % preventive expenditure stratified by dimensions 2004-2013.

## DISCUSSION

The aim of this study was to explore whether there was a paradigm of association between the trend in spending on prevention activities and a given healthcare system model. To this end, the analysis was based on data from 22 European countries, considering the percentage of preventive spending out of total healthcare spending expressed as a compound annual rate.

In 15 out of 22 countries, an increase in preventive expenditure have been observed over the considered period (2004-2023), with a compound annual growth rate peaking at 3.7%. In the remaining 7 countries, however, there was a decrease in investment in prevention, with a compound annual growth rate reaching a low of −6.3%.

Despite the increase in preventive spending in some countries, the share remains marginal compared to total healthcare spending, amplifying the findings of Gmeinder et al., who reported the same problem in Europe for the period from 2005 to 2015 (19).

As reported in the literature, prevention is often considered an ‘easy target’ for budget cuts because its benefits are only visible in the long term and there is usually no professional or commercial pressure in its favour. For this reason, prevention is a “quiet” policy that does not enjoy the same public support as policies that support interventions in the curative sphere, which cover immediate needs shared by most citizens (3,9). This modus operandi seems to be a common trend in all countries, regardless of political orientation, probably driven by the need for immediate returns in economic terms and consensus (3,20).

Despite short-term political gains, several authors argue that cutting spending on prevention is a “false economy” (9,21,22). A systematic review attempted to quantify the impact of public health interventions delivered in high-income countries, defined as “the science and art of promoting and protecting health and well-being, preventing ill-health and prolonging life through the organised efforts of society”: the median return on investment was 14.3 to 1, and the median cost-benefit ratio was 8.3 (21).

Reducing investment in this area to achieve short-term savings could not only reduce the benefits in terms of improving quality of life (23–25), but also have ruinous consequences for society in the long term, increasing pressure on healthcare services and worsening health inequalities (9,22). In fact, it should be considered that primary or secondary prevention produces significant positive externalities: when a sufficient percentage of the population is vaccinated, the circulation of the pathogen is reduced, protecting the unvaccinated (herd immunity) as well; early diagnosis of a disease not only saves the life of the individual (private benefit), but also reduces social and health costs for the entire community, improving overall public health (3). Furthermore, a major problem with preventive care is the very high asymmetric information: an ordinary citizen/patient is unable to get full information about his/her needs, his/her health risks as well as about the resulting probability of health care needs and the expenditures which will have to be supported to a health care he will consume (26).

Regarding the temporal trend, our study also identified a peak in preventive expenditure in 2020 and 2021. The sharp contraction in 2022-2023, shared by all countries, highlights the exceptional nature of the pandemic peak, linked to emergency measures such as vaccination campaigns, testing and tracking for the management of COVID-19 (19,27). The sudden return to pre-pandemic spending levels suggests that healthcare systems have missed the opportunity to invest substantially in long-term preventive care and seem to have implemented measures to reduce their deficits induced by the crisis (3). This phenomenon had already been observed during the H1N1 pandemic in 2009, where the need to manage the so-called “swine flu” caused spikes in prevention spending, followed by immediate reductions in subsequent years (19).

The results of the regression analyses show no significant association between the actor responsible for each domain characterising the specific healthcare system and the level of expenditure allocated to prevention activities. The only exception is the dimension of regulation (societal vs statal): according to our results, this is the only factor that significantly influenced the temporal trajectory of expenditure, yet with an unexpected dynamic. A widespread theory in the literature states that National Health Service-type systems, i.e. state-regulated systems, tend to maintain higher investments in prevention (3). On the contrary, our results suggest that systems in which regulation is dominated by societal actors tend to report a faster increase in expenditure, albeit slight. The interpretation of this result is very complex; the explanations proposed below cannot be inferred from our results and should be viewed with caution One reason why SHI tends to have higher preventive expenditure may be found in different approaches to funding and implementing preventive care in comparison with state-regulated systems: the societal regulation system is a mandatory, contribution-based system providing health risk cover through exhaustive benefits packages and consumers can choose among multiple sickness funds. SHI funds, through self-government, determine their own preventive offers within a legal framework; they often use prevention programmes to compete for members and may use bonuses to incentivise individual participation, which can increase the focus on preventive services (28,29). Another hypothetical explanation could be the trend towards hybridisation of healthcare systems and the crumbling of old institutional logic (30). However, looking at Figure 2, we can see how this surge in societal regulation systems is almost concurrent with the years of the pandemic. It cannot be ruled out that the above mentioned difference is influenced by the impact of the COVID-19 pandemic and that the latter has disrupted the “classic” dynamics described above.

### Limitations and strengths

Although the findings should be interpreted with caution, this study has several strengths: firstly we updated the percentage of preventive expenditure, but we estimated the slope rate as well. This second measure may be a better indicator of healthcare system choice because the slope rate is less susceptible to confounding factors such as baseline health risk and the socio-epidemiological characteristics of the population. To our knowledge, this is the first study that examines hypothetical associations between specific types of health systems and different patterns of preventive expenditure according to Bohm’s classification for health systems.

Regression analyses of each of the three dimensions - regulation, financing, provision - on the percentage of preventive expenditure may represent independent sources of action and interest in health policy. Furthermore, although it was not possible to carry out the analysis on all OECD countries, this study conducts a comparative analysis on a large sample of European countries, allowing the phenomenon to be explored in different health and economic contexts.

A limitation of this study is that this analysis has been conducted at the preventive expenditure dimensions level and has not disaggregated expenditures by specific types (primary, secondary, tertiary); this potentially could conceal variations among countries on the content and range of services offered to citizens/patients.

Consequently, analysis at the national level must be conducted to assist policymakers in making better-informed decisions about preventive care.

### Conclusions

In conclusion, for the countries analysed, our time trend analysis shows a rather heterogeneous annual rate of expenditure on preventive medicine in the considered period, yet a widespread penalising attitude towards preventive care expenditures can be generally highlighted.

This study set out that there is no correlation between the type of healthcare system and the share of expenditure allocated to prevention activities in the countries analysed. The absence of a correlation between the type of healthcare system and spending on prevention is significant from a healthcare policy perspective: the results suggest that investment in prevention is not intrinsically determined by the organisational structure of the healthcare system, but responds to external factors, such as internal economic and social needs and political decisions. This strengthens the idea that there is a strong social and political responsibility to maintain adequate and sustained investment in prevention over time, regardless of major events affecting society (e.g. the COVID-19 pandemic) and the existing healthcare system model.

Future studies should explore the role of political and contextual determinants, focusing on the most effective communication methods for guiding healthcare policy decisions.

## Data Availability

All data produced in the present study are available upon reasonable request to the authors

## Acknowledgements

We have no known conflict of interest to disclose; this research received no specific grant from any funding agency in the public, commercial or not-for-profit sectors. Furthermore, all data are available upon request from the authors.

## Notes

### Competing Interest Statement

The authors have declared no competing interest.

### Funding Statement

This study did not receive any funding

